# Bias assessment and correction for Levin’s population attributable fraction in the presence of confounding

**DOI:** 10.1101/2023.02.02.23284941

**Authors:** John Ferguson, Alberto Alvarez, Martin Mulligan, Conor Judge, Martin O’Donnell

**Affiliations:** HRB Clinical Research Facility Galway, University of Galway, Ireland; School of Medicine, University of Galway, Ireland

**Keywords:** population attributable fraction, Levin’s formula, Miettinen’s formula, asymptotic bias, risk-factor

## Abstract

In 1953, Morton Levin introduced a simple approach to estimating population attributable fractions (PAF) depending only on risk factor prevalence and relative risk. This formula and its extensions are still in widespread use today, particularly to estimate PAF in populations where individual data is unavailable. Unfortunately, Levin’s approach is known to be asymptotically biased for the PAF when the risk factor-disease relationship is confounded even if relative risks that are correctly adjusted for confounding are used in the estimator.

An alternative estimator, first introduced by Miettinen in 1972, is unbiased for the PAF provided the true relative risk is invariant across confounder strata. However, despite its statistical superiority, Miettinen’s estimator is seldom used in practice, as its direct application requires an estimate of risk factor prevalence within disease cases rather than an estimate of risk factor prevalence in the general population.

Here we describe a simple re-expression of Miettinen’s estimand that depends on the causal relative risk, the unadjusted relative risk and the population risk factor prevalence. While this re-expression is not new, it has been underappreciated in the literature, and the associated estimator may be useful in estimating PAF in populations when individual data is unavailable provided estimated adjusted and unadjusted relative risks can be transported to the population of interest. Using the re-expressed estimand, we develop novel analytic formulae for the relative and absolute asymptotic bias in Levin’s formula, solidifying earlier work by Darrow and Steenland that used simulations to investigate this bias. We extend all results to settings with non-binary valued risk factors and continuous exposures and discuss the utility of these results in estimating PAF in practice.

## Introduction

The Population Attributable Fraction (PAF), sometimes also referred to as the Population Attributable Risk and the Excess Fraction, is a central metric in epidemiology. It is useful both to ascertain the importance of a risk factor in causing disease as well as to identify the best risk factors to target in a health intervention.

Applications of attributable fractions abound in the literature starting with Richard Doll’s work in estimating the burden of lung cancer due to smoking [1], with recent examples relating to modifiable risk factors of gout in the US [2], risk factors for depression in Brazil [3] and modifiable risk factors for cancer in Denmark [4], with these studies representing only a very small sampling of recent literature.

Intuitively, PAF represents the fraction of prevalent disease cases that might have been avoided in some population if a risk factor were not present. If we express disease prevalence in the current population as *P*(*Y* = 1), *Y* being a binary indicator for disease, and the prevalence of disease that would have been observed in the target population had the risk factor been removed as *P*(*Y*_0_ = 1), PAF can be expressed as:

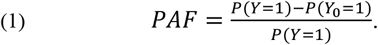

Here, we are restricting our usage of the term risk factor to apply to environmental, physiological or behavioural determinants of a particular disease; that is we do not view variables that have non-causal associations with disease as risk factors. Such factors might be binary, multi-level or continuously distributed. As an example, depending on the resolution of data capture, smoking might be coded as a binary indicator for current smoking, separate indicators for whether an individual is currently smoking, has given up smoking or has never smoked, or perhaps as total nicotine intake via smoking. Definition (1) and its interpretation, in terms of a comparison between the current population and the population had the risk factor been removed, when this is possible at least hypothetically, is valid for any type of exposure distribution. However, the details of the estimation process will differ between these three situations. We first will discuss the simplest case where the risk factor is binary before extending to more general situations.

In the binary risk factor setting, a vast literature has evolved on methods to estimate (1) with individual-level data [5]. However, it is sometimes necessary to derive estimates of PAF in scenarios where the collection of individual-level data is impossible. As an example, the Global Burden of Disease project [6] estimate population attributable fractions at a country level for differing risk factor/disease combinations. For certain countries, no individual-level data linking risk factors of interest to disease may be available. The most common approach in this situation is to substitute the estimated risk factor prevalence in the population, 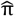, and estimated relative risk of disease (adjusted for confounders), 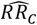, into equation (2) below. Equation (2) was introduced by Morton Levin [7], also in the context of estimating the PAF for smoking as a cause of lung cancer:

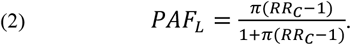

(2) is prominent throughout the literature on PAF estimation. However, as recognised by multiple authors [8],[9, 10],[11], (2) will usually differ from (1) as a quantity. In particular (2) will equal the true PAF only under the unlikely condition that the association between risk factor and outcome is unconfounded, or more technically exhibits marginal exchangeability [12]. Marginal exchangeability implies that the causal relative risk, 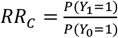, defined as the ratio of disease prevalence comparing scenarios where the entire population was exposed and the entire population was unexposed to the risk factor in question equals the unadjusted relative risk, *RR*_*U*_. See Supplementary Material Section 1B for more discussion.

### Miettenen’s definition of PAF

While (2) does not equal PAF in the absence of this ‘no confounding’ assumption, an alternative expression introduced by Miettinen [13], which involves the prevalence of the exposure within cases, *π*_*c*_, as opposed to the overall population prevalence equals equal (1) irrespective of the degree of confounding. Miettinen’s expression is given by

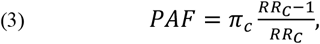

where *π*_*c*_ is now the proportion of disease cases that have the risk factor.

Equation (3) is correct provided the causal relative risk, *RR*_*C*_, is constant in differing joint strata of the confounder variables. Otherwise, a similar formula holds, replacing the causal relative risk in the overall population with the causal relative risk in individuals exposed to the risk factor (See supplementary section S2). The usual advice is to use (3) when possible [9] and not (2) as a basis for estimating PAF. However, estimates for *π* are more likely to be found in the published literature than estimates for *π*_*c*_ and as a result, researchers are more likely to use (2) in practice. For example, the Global Burden of Disease project utilise (2) rather than (3) to estimate PAF.

### A re-expression of Miettenen’s PAF

A fact that has been underappreciated in the literature is that equation (3) can be re-expressed to be a function of three quantities: the prevalence of the risk factor in the population, *π*, the causal relative risk, *RR*_*C*_, and the unadjusted relative risk, *RR*_*U*_ in the population:

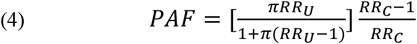

The proof that (4) equals the PAF is a relatively simple application of Bayes’ rule (supplementary section S3). This formula was known to Miettinen (see equation 8 in his 1974 pape), but has received little attention in the literature, although an equivalent formula was recently described by Susuki and Yamamoto [14]. While a somewhat simple re-expression of (3), (4) may be practically useful in estimating PAF with summary data as unadjusted relative risks are often reported together with adjusted relative risks. It is useful to notice that under no confounding, which implies *RR*_*U*_ = *RR*_*C*_, the expression (4) simplifies to Levin’s formula. This observation implies that in addition to Levin’s formula being correct under marginal exchangeability as mentioned previously, it is also correct under a slightly weaker assumption of no-confounding in risk ratio (*RR*_*U*_ = *RR*_*C*_), together with an assumption of no effect modification across confounder strata. However, formulae (2) and (4) will otherwise differ.

### Analysis of bias in Levin’s estimate

For illustration regarding both the biases at play from Levin’s approach and the potential of (4) to provide better estimates of PAF than (2) when individual-level data is unavailable, we will consider results from [15] who conducted meta-analyses to estimate the causal effect of physical inactivity on depression. Their analyses estimated both unadjusted relative risks, 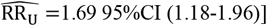, and adjusted relative risks, 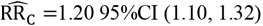, for physical inactivity. The percentage of physical inactivity (according to a certain definition) may vary greatly over differing populations, depending on factors such as culture and age structure. Let’s say that for a particular population, an estimate of the percentage of physical inactivity is *π* = 0.5. Plugging in the estimated adjusted relative risk into Levin’s formula results in an estimated PAF of 9.1%, whereas using (4), the correctly estimated PAF is 10.5%. Suppose we wish instead to estimate the PAF in a more active population with an estimate of physical inactivity: *π* = 0.2. In this case, the bias is smaller but the relative bias is larger: Levin’s formula gives an estimate of 3.8% whereas (4) generates an estimate of 5.0%. These are examples of the general behaviour we would expect of the bias as we will show below.

[10] investigated the bias of Levin’s formula using simulation, and showed that as the degree of confounding, specified by *max*{*C*, 1/*C*} where 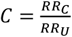 gets larger, with no confounding in risk ratio being represented by *C* = 1, the magnitude of *relative* bias in Levin’s formula will increase. When *C* < 1, one would expect that *PAF*_*L*_ < *PAF* (as is the case in the effect of physical inactivity on depression). Their simulations also indicate that this relative bias will be larger for smaller risk factor prevalences, as again observed in the previous example. However, having an explicit formula for relative bias allows a more rigorous analysis of limiting behaviour for extreme values of *C* than was possible in [10].

Given the formula (4), an expression for the relative asymptotic bias can be trivially derived by simply taking the ratio of (3) and (4). After some simplification (supplementary section S4) this results in:

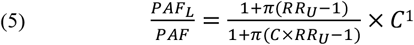

Observing that the partial derivative 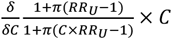 is strictly positive when fixing *RR*_*U*_ and *π* (Supplementary Section S8) it follows that (5) is an increasing function of *C*, fixing values of these variables. Given that (5) equals 1 at *C* = 1 (that is Levin’s formula is unbiased when there is no confounding in risk ratio) this indicates that the degree of relative bias gets larger as the degree of confounding increases in either direction *C* > 1: *C* ↑ ∞ or *C* < 1: *C* ↓ 0. It is useful to analyse the relative (and absolute bias) separately in these two scenarios *C* > 1 and *C* < 1. See Supplementary Section S8 for more detail regarding these analyses. A graphical representation of the associated biases (similar to the plots in [10], but with identification of limiting biases and behaviour of Levin’s formula using *RR*_*U*_ in place of *RR*_*C*_ is given by Figure 1).

**Figure 1:**
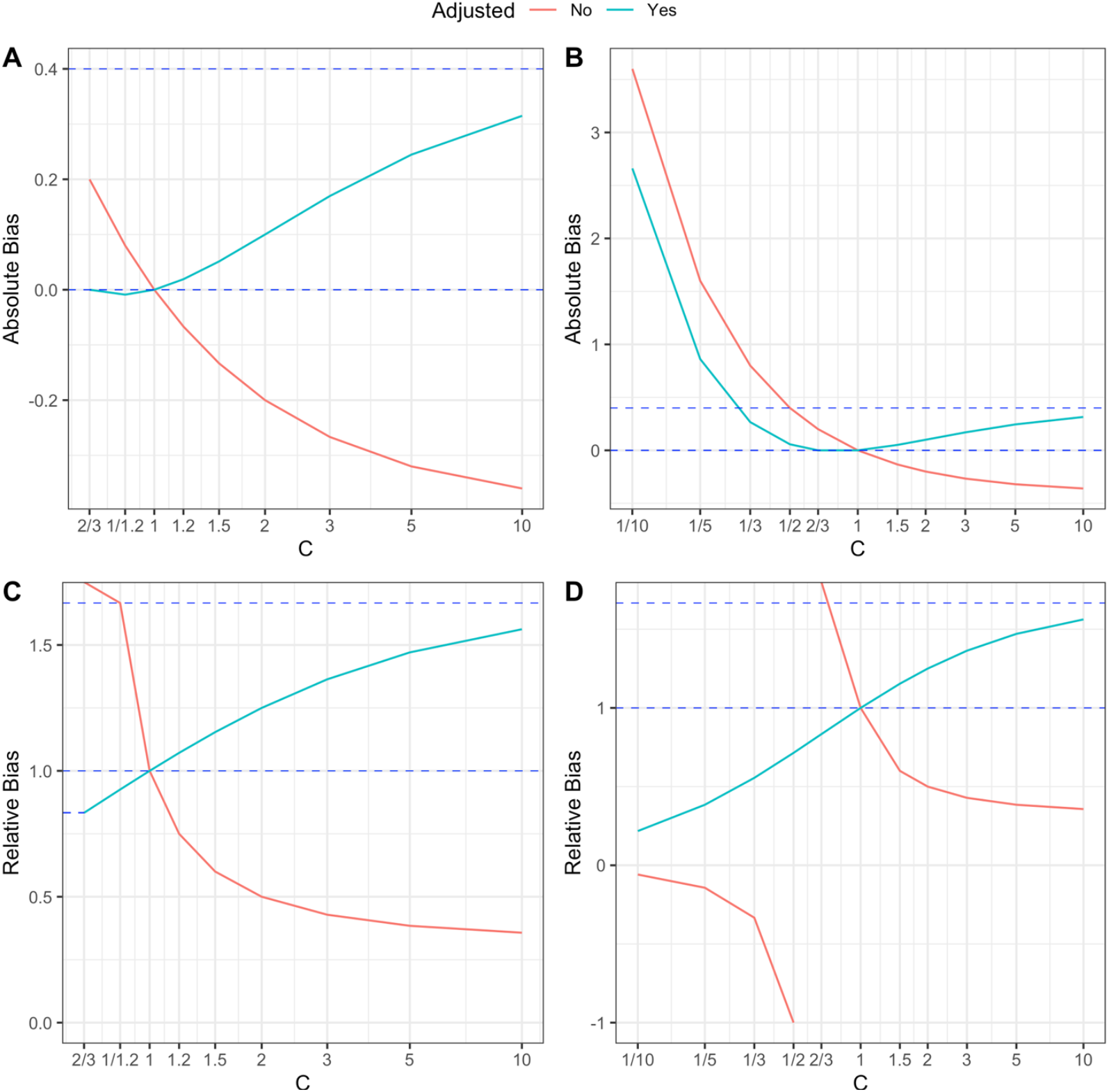
Plots for absolute and relative error in Levin’s formula when *RR*_*U*_ = 1.5 and *π* = 0.5 over a range for the confounding ratio *C*. A: absolute bias, *RR*_*C*_ ≥ 1, B: absolute bias, no restriction on *RR*_*C*_, C: relative bias, *RR*_*C*_ ≥ 1, D: relative bias, no restriction on *RR*_*C*_. In the attached plots, the bias for the adjusted version, 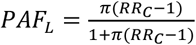of Levin’s formula is given in blue, while biases for the unadjusted version, 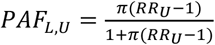 are given in red. The upper blue horizontal lines are the limiting absolute bias: 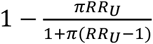 and relative bias: 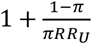 for *PAF*_*L*_ as *C* → ∞. Note that the absolute bias of *PAF*_*L*_ is 0 at both *C* = 1/*RR*_*U*_ and *C* = 1,while the relative bias converges to 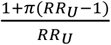 as *C* approaches 1/*RR*_*U*_ (as tagged on the y-axis of plot C). The absolute and relative biases of the unadjusted version of Levin’s formula are more erratic. In particular, the relative bias function is negative when *RR*_*C*_ < 1 or equivalently 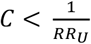 indicating that the *PAF*_*L,U*_ is suggesting an incorrect direction of association, *PAF*_*L*_ is always larger than 0 for all *C* > 0 and converges to 0 as *C* → 0.

### Bias in *PAF*_*L*_ when *C* > 1

For values of *C* > 1, Levin’s formula is biased above with the relative bias increasing toward a limit of 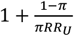 as *C* → ∞ (note again that this analysis assumes *π* and *RR*_*U*_ are fixed). As one would expect under no confounding in risk ratio: 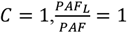 (that is the Levin estimand and true PAF are equal). Provided *RR*_*U*_ > 1, the absolute bias: *PAF*_*L*_ − *PAF* also increases as *C* increases (from *PAF* − *PAF* = 0 when *C* = 1 to 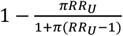 as *C* → ∞). Note that Levin’s formula, (2), converges to 1 as *C* → ∞ irrespective of the values of *π* and *RR*_*U*_, whereas the true *PAF* converges to 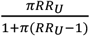 as *C* gets very large. In practice, imagining what the bias might be for extremely large values of *C* is interesting theoretically in it demonstrates the maximum possible bias in Levin’s approach; but whether one would see such extreme confounding in a real example is debatable. For instance, a scenario where extreme values of *C* are possible is if there is a second risk factor, *X*^∗^ that acts as a confounder for the relationship *X* → *Y*, with a strong causal influence on *X* so much so that *X*^∗^ ∼ *X* in the population, and such that the direct effect of *X*^∗^ not mediated through *X* is to strongly reduce the likelihood of disease (by a factor of 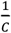), but the effect of *X* on Y is to strongly increase the likelihood of disease by a factor of *C*. Such scenarios are likely implausible in practical situations.

When 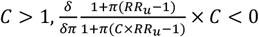 (fixing *RR*_*U*_ and *C*), indicating that as risk factor prevalence, *π*, decreases relative bias, (5), increases. Given that relative bias is larger than 1 when *C* > 1, this indicates that relative bias worsens as risk factor prevalence gets smaller, although the situation for the degree of absolute bias is more complicated (absolute bias will converge to 0 as *π* ↓ 0 and as *π* ↑ 1).

### Bias in *PAF*_*L*_ when *C* ≤ 1

For *C* < 1, Levin’s formula is biased below and the relative bias becomes progressively worse, eventually converging towards 0 as *C* ↓ 0 (a realm in which risk factors are protective and attributable fractions are negative). However, we usually are more interested in risk factor codings where *RR*_*C*_ ≥ 1. Assuming that *RR*_*U*_ > 1, the absolute bias *PAF*_*L*_ − *PAF* is 0 under no causal effect, *RR*_*C*_ = 1, (or equivalently for 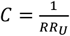) in addition to being 0 when *C* = 1 (that is under no confounding). By continuity of the expressions (2), (3) and (4) as functions of *C* we can argue that when the true causal effect is very small, *RR*_*C*_ ≈ 1, the absolute bias in Levin’s formula will be negligible, while the relative bias will be approximately 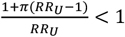, although arguably the relative bias is not so important in this case. These results imply that Levin’s formula will be appropriate in scenarios where the true causal effect is very small or where there is very little confounding.

When 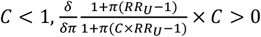 (fixing *RR*_*U*_ and *C*), indicating that as risk factor prevalence decreases, the relative bias function decreases. Given that the relative bias function is smaller than 1 when *C* < 1, this indicates again that relative bias gets worse as risk factor prevalence gets smaller, although as in the situation where *C* > 1 absolute bias converges to 0 as *π* converges to 0 or as *π* converges to 1.

### PAF for exposures with more general distributions

When the distribution of exposure is multi-category or continuous, PAF can be defined as the fraction of prevalent (or incident) disease cases that would have been avoided in a population where the value of the exposure was fixed at a particular value (or within a range of values) that minimises the probability of disease (this value is known as the minimum risk exposure value or MREV). While with individual-level data it is possible to estimate the MREV [16], when deriving estimates of PAF from summary estimates one usually assumes a pre-determined MREV [6]) For example, in the simple case of an exposure such as air pollution which can be eliminated, the MREV would be presumed to be 0 (that is elimination), with non-zero values being appropriate for exposures like blood pressure or sodium consumption. The causal relative risk is now a function, *RR*_*C*_ (*x*) comparing the increased prevalence of disease if the population were all exposed to exposure level *x* relative to disease prevalence if the same were all exposed to the MREV. Under the assumption that for each possible exposure value, *x, RR*_*C*_ (*x*) is constant within confounder strata, Miettinen’s formula (4) can be extended as follows:

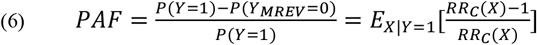

where *E*_*X*|*Y*=1_[*f*(*X*)] denotes the average of a function, *f*(*X*), of the exposure, *X* within the population of individuals with disease (supplementary section S5). Note that this formula is valid for both continuous exposures (such as air pollution) and multi-category exposures (such as a three-level coding of smoking in terms of current smokers/former smokers and never-smokers), and simplifies to (3) in the setting of a binary exposure.

As shown in supplementary section S7, (6) can be re-expressed using Bayes’ Rule as follows:

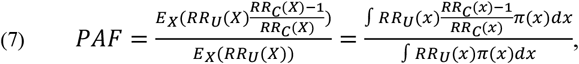

with *R*_*U*_(*x*) representing the unadjusted relative risk function that compares the prevalence of disease in the strata of the population exposed to the value *x* of the risk factor with the prevalence in the strata exposed to the MREV, and *E*_*X*_ is the population expectation operator. The right-hand side of (7) indicates that one can calculate PAF by numerical integration of the quantities 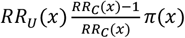 and *RR*_*U*_ (*x*)*π*(*x*) (here we are assuming that the exposure distribution is absolutely continuous with density *π*(*x*)). In the special case that the exposure is discrete, but with more than two levels: *X* ∈ {0,1, …, *L*}, with the population prevalences of each level: *π*(0), …, *π*(*L*), and MREV=0, equation (7) can be re-expressed as:

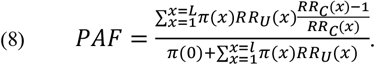

### Bias from Levin’s approach for more general exposure distributions

The generalisation of Levin’s formula to continuous exposure distributions is given by:

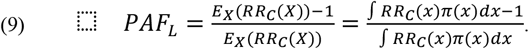

The formula for multi-category exposures is very almost identical, apart from replacing the integral in 9 with a summation over the values, *x*, that the exposure can take. See Table 1 for a comparison of Levin’s and Miettinen’s approach for the binary, multi-category and continuous exposures. Comparing equations (7) and (9), it follows immediately that under no confounding, that is *RR*_*U*_ (*x*) = *RR*_*C*_(*x*) for every exposure value *x*, Levin’s formula is again unbiased. However, analysis of bias in Levin’s formula is more complicated when the distribution of the exposure is non-binary, although under certain special settings, one can analyse bias in the same way as in the binary exposure case. For instance, suppose that the *MREV* = 0 and the prevalence of the minimum risk exposure level in the general population is 1 − *π* where 1 > 1 − *π* > 0. Assuming that the confounding ratio 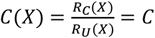 at all exposure levels not equal to the MREV, the relative bias in the Levin estimate is:

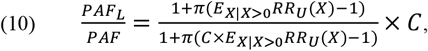

**Table 1.** Below, we show the identification formulae for PAF and PAF_*L*_, for binary, multicategory and continuous exposure distributions, under the assumption of no effect modification. *π* represents the population prevalence of a binary risk/ factor and *π*_*C*_ the prevalence within disease cases, with similar notation representing probability mass functions and density functions when the exposure is multi-category or continuously distributed. An implicit assumption for the multicategory and continuous formulae is that the MREV (minimum risk exposure value) is equal to 0. In all settings, PAF is expressed using the traditional Miettinen formula (in terms of *π*_*C*_ and RR_*C*_) and the 3 variable formula advocated in this manuscript which can be used if estimates of both the unadjusted and adjusted relative risks are available.

| | $PAF_L$ | $PAF$ | $PAF$ |
| --- | --- | --- | --- |
| | | (in terms of $\pi_c$ and $RR_C$) | (in terms of $\pi$, $RR_U$ and $RR_C$) |
| **Binary risk factor** | $\frac{\pi(RR_C - 1)}{1 + \pi(RR_C - 1)}$ | $\pi_c \frac{RR_C - 1}{RR_C}$ | $\left[ \frac{\pi RR_U}{1 + \pi(RR_U - 1)} \right] \frac{RR_C - 1}{RR_C}$ |
| **Multicategory risk factor** | $\frac{\sum_{x=1}^L \pi(x)(RR_C(x) - 1)}{1 + \sum_{x=1}^L \pi(x)(RR_C(x) - 1)}$ | $\sum_{l=1}^L \pi_c(x) \frac{RR_C(x) - 1}{RR_C(x)}$ | $\frac{\sum_{x=1}^L \pi(x) RR_U(x) \frac{RR_C(x) - 1}{RR_C(x)}}{\pi(0) + \sum_{x=1}^L \pi(x) RR_U(x)}$ |
| **Continuous risk factor** | $\frac{\int RR_C(x) \pi(x) dx - 1}{\int RR_C(x) \pi(x) dx}$ | $\int \frac{RR_C(x) - 1}{RR_C(x)} \pi_c(x) dx$ | $\frac{\int RR_U(x) \frac{RR_C(x) - 1}{RR_C(x)} \pi(x) dx}{\int RR_U(x) \pi(x) dx}$ |

, an expression very similar to (5) and which indicates that larger relative biases are expected under larger levels of confounding (that is *C* << 1 or *C* >> 1). For continuous exposures, the assumption that a non-zero proportion of the population is exposed to the minimum exposure level may well be implausible. However, a similar equation to (10) will hold appropriately if there is a range of exposure values, *R*, having non zero probability in the population which approximately minimise counterfactual risk: 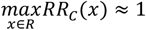, where 0 < *π* = *P*(*X* ∉ *R*) < 1 and 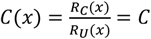 when *x* ∉ *R*. The modified equation then would be:

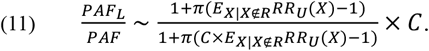

The proofs of equations (10) and (11) are given as supplementary section S9.

## Discussion

We use this final section mostly to discuss caveats and limitations to our suggested approaches. First, we have suggested that where possible equation (4) should be used instead of Levin’s formula, (2), to estimate PAF when individual-level data is unavailable. Equation (4) requires estimates *RR*_*U*_ and *RR*_*C*_ of the unadjusted and causal relative risks. To correctly estimate PAF these estimates need to be transportable to the target population. While transportability of the causal relative risk is an issue both for equation (4) and Levin’s approach, (2), transportability of the unadjusted relative risk is an additional assumption and there is no good reason to expect it to hold. Fortunately, often plugging in an incorrect estimated unadjusted relative risk into (4) will represent an improvement over Levin’s formula (3): provided the patterns of confounding in the source and target population are reasonably similar, one might expect *RR*_*U*_ and *RR*_*U*_ to be reasonably similar and equation (4) to be more accurate than equation (2).

As noted by other authors, the bias using an adjusted relative risk with Levin’s formula is generally quite small [10] and likely insubstantial compared to other biases involved in estimating a PAF. For instance, in the example regarding the relationship between physical inactivity and depression, there are additional questions about the consistency of ascertainment and measurement of both the exposure (inactivity) and outcome (depression). The prevalence of defined inactivity (and the associated relative risk) will change depending on what the investigator considers to constitute inactivity, and the definition may differ across different studies. Similar issues arise in the consistency of ascertainment and measurement of depression. These issues reduce the real-world meaning and actionability of any estimated PAF. On a related point, physical activity should really be measured on a continuum; a binary definition of inactivity will likely underestimate associated disease burden. For instance, a better approach to calculating PAF might determine an optimal level of physical activity (that perhaps differs dependent on age and other characteristics of a person) and estimate disease prevalence in a hypothetical population where everybody had at least this optimal level of activity; however, implementation of such an estimator may be practically challenging. Finally, the assumption that the causal relative risk is constant across differing confounding strata will usually be dubious (individuals exposed or not exposed to differing patterns of confounding variables have differing baseline probabilities of disease making the same relative effect of a risk factor unlikely). If the causal relative risk varies over differing strata of confounders, formulae for *PAF* for binary and general exposure distributions are as follows:

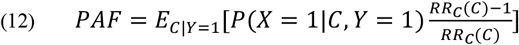

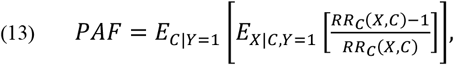

where *RR*_*C*_(*c*) is regarded as the causal relative risk within confounder stratum *C* = *c* in (12) and *RR*_*C*_(*x, c*) as the causal relative risk comparing exposure levels *x* and the MREV in confounder stratum *C* = *c* in (13). Note that (12) can be re-expressed as: 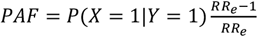, where 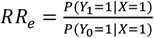 is the causal relative risk restricted to the population exposed to the risk factor, as Miettinen originally showed in 1972. These formulae are derived in supplementary sections S2 and S6 and are equivalent to, and extend of in the case of (13), the formulae suggested in [17], and without a no-effect modification assumption require individual-level data (incorporating effect modification between risk factors and confounders) to estimate. When relative risks vary over confounder strata, the marginal causal relative risk, *RR*_*C*_ (defined as the ratio of disease probabilities if everyone was exposed and everyone was exposed to the risk factor), and the causal relative risk in exposed individuals are both weighted average of confounder-strata specific causal relative risks, *RR*_*C*_(*c*) (see Supplementary Appendix S10). As a result, one would not expect large differences between (4) and (12) under moderate levels of effect modification.

While equivalent results to equation (4) exist for multi-category and continuous exposures (equations (7)-(8)) these may be not as useful in practice as equation (4) as they require specification of unadjusted relative risks comparing many levels of exposure to baseline. As a result, Levin’s formula (despite its bias) may forever be the method of choice for estimating PAFs and impact fractions for continuous exposures with summary data. However, the biases in Levin’s formulae are often dismissed in this setting. If the formula is to be used it is important to recognise its bias and have some awareness of the likely extent of the bias. In this regard, if there is a range of values of the exposure which approximately minimises risk and it’s plausible that the confounding parameter: 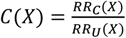 is approximately constant outside of this range, equation (11) may be useful in determining the likely error from using Levin’s approach.

Here we advocate that authors use equation (4) when estimates of unadjusted and adjusted relative risks are available. In contrast, in the literature we’ve noticed several examples of Levin’s formula being applied with unadjusted relative risks in observational settings [18], [19]. While this will generate correct results under no confounding, we would not advise this practice. First, if there is no confounding, either an adjusted or unadjusted relative risk would be appropriate to use in Levin’s approach, that is both should generate statistically consistent estimates of the true causal relative risk, provided no effect modifiers are included in the set of variables that are adjusted for in the adjusted relative risk. Second, in observational settings, there usually will be some degree of confounding in which case substituting unadjusted relative risks into Levin’s formula will often result in more egregious bias than if appropriately adjusted relative risks were instead used in the same formula. For example, using Levin’s formula with an unadjusted relative will ‘flip’ the direction of the putative PAF when *RR*_*U*_ < 1 and *RR*_*C*_ > 1, so that while *PAF* > 0 (the risk factor causes disease), 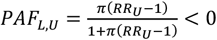, that is the risk factor would seem to be protective when using an unadjusted relative risk in Levin’s approach. In contrast, this problem does not happen with the true causal relative risk: 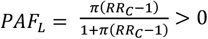. Plots B and D in Figure 1 show that, as one might expect, Levin’s formula also flips the direction of the causal effect if *RR*_*U*_ > 1 and *RR*_*C*_ < 1. In addition, for very small causal effects with *RR*_*C*_ ≈ 1, *PAF*_*L*_ ≈ 0 irrespective of the value of the confounding ratio *C*, however in the same scenario *PAF*_*L, U*_ can be quite biased if *C* is very large or very small. Going back to the example involving physical inactivity and depression discussed earlier, using the unadjusted relative risk estimate, 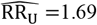, in Levin’s formula generates an estimated PAF of 25.7% which is likely a huge overestimate compared to the estimate of 9.0% from using the adjusted estimate, 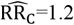. Note that using equation (4) results in an estimate of 10.5% in that example.

## Statements and Declarations

### Data Availability

All of the results in this manuscript are proven in the supplementary pdf. Confidence intervals for PAF_N_ and PAF, under the assumptions of independent estimates of RR_U_, RR_C_ and π are available through the R package graphPAF downloadable from the GitHub repository: github.com/johnfergusonNUIG/graphPAF.

## Footnotes

1 Note that (5) is technically the relative difference (or error) between the estimands *PAF*_*L*_ and *PAF*. However, if statistically consistent estimates of *RR*_*C*_ and *π* are plugged in to Levin’s formula, (5) will represent the asymptotic bias of the resulting estimator. As a result, we will (somewhat informally) refer to (5) and comparable quantities as biases throughout the manuscript.

