## Supplementary material for "Bias assessment and correction for Levin’s population attributable fraction in the presence of confounding"

**S1A** Proof that $PAF = \pi_c \frac{RR_C - 1}{RR_C}$, where $\pi_c = P(X = 1 \mid Y = 1)$ and $RR_C = \frac{P(Y_1 = 1)}{P(Y_0 = 1)}$ provided that:

1) Conditional exchangeability: $Y_x \perp X \mid C = c$, holds within each confounder strata $c$, for $x \in \{0,1\}$[1]
2) Consistency: If $X = x$, $Y = Y_x$, for $x \in \{0,1\}$
3) $RR_C(c) = \frac{P(Y = 1 \mid X = 1,c)}{P(Y = 1 \mid X = 0,c)} = K$[2] is constant over confounder strata $C = c$

**Proof**

First, we note that:

$$P(Y = 1 \mid X = x, c) = P(Y_x = 1 \mid X = 1,c) = P(Y_x = 1 \mid c), x \in \{0,1\}$$

with the first equality due to consistency and the second due to assumption 1) above. This indicates that:

$$RR_C(c) = \frac{P(Y = 1 \mid X = 1,c)}{P(Y = 1 \mid X = 0,c)} = \frac{P(Y_1 = 1 \mid c)}{P(Y_0 = 1 \mid c)} = K$$

that is the ratio of conditional probabilities of disease under exposure and non-exposure in stratum $C = c$ equates to the causal relative risk in stratum $c$, which we assume is a constant $K$

Now note that

---

[1] Technically 'mean' conditional exchangeability, that is $P(Y_a^* = 1 \mid X = a, C = c) = P(Y_a^* = 1 \mid C = c)$ for any values $\{a, a^*\} \in \{0,1\}^2$ is sufficient

[2] Note, in the proof below and subsequent proofs, we utilise upper case letters to represent random variables and lower case letters to represent values those random variables may take. We assume for simplicity that the distribution of confounders is discrete, so that we can represent expectations of functions of confounder variables over the population: $E_C f(C)$ as a sum over differing values $\sum_c f(c)P(c)$, with $P(c)$ is shorthand for $P(C = c)$. We also abbreviate conditional expectations of confounder variables such as: $P(X = 1 \mid C = c)$ as $P(X = 1 \mid c)$.

$$RR_C = \frac{P(Y_1 = 1)}{P(Y_0 = 1)} = \frac{E(P(Y_1 = 1 | C))}{E(P(Y_0 = 1 | C))} = \frac{E(P(Y_0 = 1 | C)\frac{P(Y_1 = 1 | C)}{P(Y_0 = 1 | C)})}{E(P(Y_0 = 1 | C))} = K$$

indicating that $RR_C(c) = RR_C$ for all confounder strata $c$

The proof proceeds as follows:

$$PAF = \frac{P(Y = 1) - P(Y_0 = 1)}{P(Y = 1)}$$

$$= \frac{P(Y = 1) - E_C(P(Y_0 = 1 | C))}{P(Y = 1)}$$

$$= \frac{P(Y = 1) - E_C(P(Y_0 = 1 | X = 0, C))}{P(Y = 1)}$$ (by assumption 1)

$$= \frac{P(Y = 1) - E_C(P(Y = 1 | X = 0, C))}{P(Y = 1)}$$ (by consistency)

$$= \frac{E_{X,C}(P(Y = 1 | X, C)) - E_C(P(Y = 1 | X = 0, C))}{P(Y = 1)}$$

$$= \frac{E_C(P(X = 1 | C)P(Y = 1 | X = 1, C) + P(X = 0 | C)P(Y = 1 | X = 0, C)) - E_C(P(Y = 1 | X = 0, C))}{P(Y = 1)}$$

$$= \frac{E_C(P(X = 1 | C)P(Y = 1 | X = 1, C) + P(X = 0 | C)P(Y = 1 | X = 0, C) - P(Y = 1 | X = 0, C))}{P(Y = 1)}$$

$$= \frac{E_C(P(X = 1 | C)P(Y = 1 | X = 1, C) - P(X = 1 | C)P(Y = 1 | X = 0, C))}{P(Y = 1)} *$$

$$= (RR_C - 1)\frac{E_C(P(X = 1 | C)P(Y = 1 | X = 0, C))}{P(Y = 1)}$$ (since $RR_C = \frac{P(Y = 1 | X = 1, C)}{P(Y = 1 | X = 0, C)}$)

$$= \frac{(RR_C - 1)}{RR_C} \frac{E_C(P(X = 1 | C)P(Y = 1 | X = 1, C))}{P(Y = 1)}$$ (since $RR_C^{-1} = \frac{P(Y = 1 | X = 0, C)}{P(Y = 1 | X = 1, C)}$)

$$= \frac{P(X = 1, Y = 1)}{P(Y = 1)} \times \frac{RR_C - 1}{RR_C}$$ (since $P(X = 1, Y = 1) = E_C(P(X = 1, Y = 1 | C))$)

$$= P(X = 1 | Y = 1) \times \frac{RR_C - 1}{RR_C}$$

$$= \pi_c \frac{RR_C - 1}{RR_C}$$

**S1B.** Proof that $PAF = \pi \frac{RR_U - 1}{1 + \pi(RR_U - 1)}$ under

1) Marginal exchangeability: $Y_x \perp X$ for $x \in \{0,1\}$
2) Consistency: If $X = x$, $Y = Y_x$, for $x \in \{0,1\}$

**Proof**

Under the above marginal exchangeability assumption, one can show equality of the causal and unadjusted relative risks:

$$RR_C = \frac{P(Y_1 = 1)}{P(Y_0 = 1)} = \frac{P(Y_1 = 1 | X = 1)}{P(Y_0 = 1 | X = 0)} = \frac{P(Y = 1 | X = 1)}{P(Y = 1 | X = 0)} = RR_U$$

where assumption 1 is necessary for the first equality and assumption 2 for the second equality.

$$PAF = \frac{P(Y = 1) - P(Y_0 = 1)}{P(Y = 1)} = \frac{P(Y = 1) - P(Y = 1 | X = 0)}{P(Y = 1)} \text{ (assumptions 1 and 2)}$$

$$= \frac{P(Y = 1 | X = 0)P(X = 0) + P(Y = 1 | X = 1)P(X = 1) - P(Y = 1 | X = 0)}{P(Y = 1)}$$

$$= \frac{P(X = 1)(P(Y = 1 | X = 1) - P(Y = 1 | X = 0))}{P(Y = 1)}$$

$$= \frac{P(X = 1)P(Y = 1 | X = 1)(1 - RR_U^{-1})}{P(Y = 1)} = \pi_c(1 - RR_U^{-1})$$

An application of Bayes' Rule (which here will demonstrate that: $\pi_c = \frac{\pi RR_U}{1 + \pi(RR_U - 1)}$, see Section 3 of the Supplementary material) then shows that the above expression equals:

$$PAF = \frac{\pi RR_U}{1 + \pi(RR_U - 1)}(1 - RR_U^{-1}) = \frac{\pi(RR_U - 1)}{1 + \pi(RR_U - 1)}$$

Since we have also shown $RR_U = RR_C$, this can also be expressed as:

$$PAF = \frac{\pi(RR_C - 1)}{1 + \pi(RR_C - 1)}$$

Note that no assumptions regarding effect modification were necessary in the above derivation.

**Section 2**

**Expressions for Miettinen's formula under effect modification**

In this section, we give proofs of more general formulations of Miettinen's formula. We assume 1) and 2) from Section S1, but now allow the causal relative risk: $RR_C(c)$ to vary over covariate strata $c$.

**S2A Expression for Miettenin's PAF when the causal relative risk $RR_C(c)$ varies over confounder strata:**

First we show that:

$$PAF = E_{C|Y=1}(P(X=1 \mid Y=1, C)\frac{RR_C(C)-1}{RR_C(C)})$$

where $RR_C(c) = \dfrac{P(Y_1 = 1 \mid C = c)}{P(Y_0 = 1 \mid C = c)}$ is the causal relative risk within stratum $C = c$.

**Proof:**

The steps to show this result are identical to as in Supplementary Section 1A, up to the asterisk in that proof. The proof proceeds as follows

$$PAF = \frac{E_C(P(Y=1, X=1 \mid C)\frac{RR_C(C)-1}{RR_C(C)})}{P(Y=1)}$$

$$= \frac{E_C(P(Y=1 \mid C)P(X=1 \mid Y=1, C)\frac{RR_C(C)-1}{RR_C(C)})}{P(Y=1)}$$

$$= \frac{\sum_c P(c)P(Y=1 \mid C)P(X=1 \mid Y=1, C)\frac{RR(c)-1}{RR(c)}}{P(Y=1)}$$

$$= \sum_c P(c \mid Y=1)P(X=1 \mid Y=1, C)\frac{RR(c)-1}{RR(c)}$$

$$= E_{C|Y=1}(P(X=1 \mid Y=1, C)\frac{RR_C(C)-1}{RR_C(C)}) \qquad (1)$$

When individual data $(c_i, x_i, y_i)_{i \leq N}$ is available from the population of interest, the above formula: $PAF = E_{C|Y=1}(P(X=1|Y=1,C)\frac{RR_C(C)-1}{RR_C(C)})$ can re be used to estimate PAF by averaging $I\{x_i = 1\}\frac{\hat{RR}_C(c_i)-1}{\hat{RR}_C(c_i)}$ over the subset of cases in the data: $\{i \leq N : y_i = 1\}$ where $\hat{RR}_C(c_i)$ is the estimated relative risk of disease for an individual with covariates $c_i$. Regression type methods can facilitate estimation of $\hat{RR}_C(c_i)$.

**S2B Original formulation for Miettinen's formula:**

Here we will show that:

$$PAF = P(X=1|Y=1)\frac{RR_e - 1}{RR_e}$$

where $RR_e$ is the causal relate risk: $\frac{P(Y_1 = 1|X=1)}{P(Y_0 = 1|X=1)}$ in the subpopulation exposed to the risk factor.

**Proof**

We start with the expression for PAF that was proven in S2A

$$PAF = E_{C|Y=1}(P(X=1|Y=1,C)\frac{RR_C(C)-1}{RR_C(C)})$$

$$= \sum_c P(c|Y=1)P(X=1|Y=1,c)\frac{RR_C(c)-1}{RR_C(c)} \qquad (2)$$

Since

$$P(X=1|Y=1,c) = P(Y=1)P(X=1|Y=1)P(c|X=1,Y=1)/P(c)P(Y=1|c)$$

and $P(c \mid Y=1) = P(c)P(Y=1 \mid c)/P(Y=1)$, (2) equals:

$$P(X=1 \mid Y=1) \sum_c P(c \mid X=1, Y=1) \frac{RR_C(c)-1}{RR_C(c)} \tag{3}$$

Noting $\frac{RR_c(c)-1}{RR_c(c)} = \frac{P(Y=1 \mid X=1, c) - P(Y=1 \mid X=0, c)}{P(Y=1 \mid X=1, c)}$ and

$p(c \mid X=1, Y=1) = P(Y=1 \mid X=1, c)P(c)P(X=1 \mid c)/P(X=1)P(Y=1 \mid X=1)$,
it follows that the above display is equal to

$$P(X=1 \mid Y=1) \sum_c \frac{P(c)P(X=1 \mid c)[P(Y=1 \mid X=1, c) - P(Y=1 \mid X=0, c)]}{P(X=1)P(Y=1 \mid X=1)} \tag{3}$$

Noting:
$P(c)P(X=1 \mid c)/\{P(X=1)P(Y=1 \mid X=1)\} = p(c \mid X=1)/P(Y=1 \mid X=1)$

and $\sum_c P(c \mid X=1)P(Y=1 \mid X=1, c) = P(Y=1 \mid X=1)$, the above equals:

$$P(X=1 \mid Y=1) \frac{P(Y=1 \mid X=1, c) - \sum_c P(c \mid X=1)P(Y=1 \mid X=0, c)}{P(Y=1 \mid X=1)} \tag{4}$$

Finally, letting the causal relative risk in risk in the exposed be:
$RR_e = \frac{P(Y_1=1 \mid X=1)}{P(Y_0=1 \mid X=1)} = \frac{P(Y=1 \mid X=1)}{E_{C|X=1}P(Y_0=1 \mid X=1, C)} = \frac{P(Y=1 \mid X=1)}{E_{C|X=1}P(Y=1 \mid X=0, C)}$

where the first equality assumes consistency and the second equality assumes conditional exchangeability and consistency , it follows that in general PAF can be expressed as:

$$P(X=1 \mid Y=1) \frac{RR_e - 1}{RR_e} \tag{5}$$

**S3. Proof that under the assumptions of S1,** $PAF = \frac{\pi RR_U}{1+\pi(RR_U-1)}\frac{RR_C-1}{RR_C}$

By S1A, the result will be true if $\pi_c = \frac{\pi RR_U}{1+\pi(RR_U-1)}$; this can be proven using Bayes' Rule.

**Proof:**

$$
\begin{aligned}
\pi_c &= P(X=1\,|\,Y=1)\\
&= \frac{P(X=1,Y=1)}{P(Y=1)}\\
&= \pi\frac{P(Y=1\,|\,X=1)}{\pi P(Y=1\,|\,X=1)+(1-\pi)P(Y=1\,|\,X=0)}\\
&= \pi\frac{1}{\pi+(1-\pi)RR_U^{-1}}\\
&= \frac{\pi RR_U}{1+\pi(RR_U-1)}
\end{aligned}
$$

This implies that:

$$PAF = [\frac{\pi RR_U}{1+\pi(RR_U-1)}]\frac{RR_C-1}{RR_C}$$

**S4. Derivation of Formula (5) in the main manuscript for relative bias under the assumptions in S1**

Remembering that $PAF_L = \pi \frac{RR_C - 1}{1 + \pi(RR_C - 1)}$ and under the assumptions in S1,

$PAF = [\frac{\pi RR_U}{1 + \pi(RR_U - 1)}]\frac{RR_C - 1}{RR_C}$, it follows that:

$$B = \frac{PAF_L}{PAF}$$

$$= \frac{(RR_C - 1)(1 + \pi(RR_U - 1))}{RR_U(1 + \pi(RR_C - 1))} \times \frac{RR_C}{RR_C - 1}$$

$$= \frac{1 + \pi(RR_U - 1)}{1 + \pi(RR_C - 1)} \times \frac{RR_C}{RR_U}$$

$$= \frac{1 + \pi(RR_U - 1)}{1 + \pi(C \times RR_U - 1)} \times C$$

**S5. Generalisation of Miettinen's formula for continuous (or multi-category) exposure distributions**

For a strata of confounders, $C = c$ we will assume that $RR(x,c) = \dfrac{P(Y=1 \mid X=x, C=c)}{P(Y=1 \mid X=0, C=c)} = K_x$ is constant over all confounder strata $C = c$ for each exposure value $x$.

We assume without loss of generality, that the Minimum risk exposure value is 0 (MREV = 0), and write, $RR_C(x) = \dfrac{P(Y_x = 1)}{P(Y_0 = 1)}$ for the causal relative risk at exposure value $X = x$, relative to the MREV. Note this is somewhat an abuse of notation since in section S3, we used $RR_C(c)$ to refer to the causal relative risk in confounder stratum $C = c$. In practice, we use context to distinguish $RR_C(c)$ and $RR_C(x)$, with the convention that we always use fixed $c$ to represent confounder strata and fixed $x$ to represent an exposure value.

Below, we also assume conditions 1 and 2 (conditional exchangeability and consistency) given in S1, but now relative to the continuously distributed exposure $X$; previously the implicit assumption was a binary exposure.

Finally, note the symbol $\pi$ is used to denote a generic density function for the exposure $X$, with its arguments determining the precise density or conditional specified.

**Proof**

Note similarly to in S1:

$$RR(c,x) = \frac{P(Y=1 \mid X=x, c)}{P(Y=1 \mid X=0, c)} = \frac{P(Y_1 = x \mid c)}{P(Y_0 = 1 \mid c)} = K_x$$

where the second equality follows by using both consistency and conditional exchangeability assumptions/

which implies that, for each fixed value $x$

$$RR_C(x) = \frac{P(Y_1 = 1 | X = x)}{P(Y_0 = 1 | X = x)} = \frac{E_C(P(Y_1 = 1 | C, X = x))}{E_C(P(Y_0 = 1 | C, X = x))}$$

$$= \frac{E_C(P(Y_0 = 1 | C, X = x)\frac{P(Y_1 = 1 | C, X = x)}{P(Y_0 = 1 | C, X = x)})}{E_C(P(Y_0 = 1 | C, X = x))} = K_x$$

so that $\dfrac{P(Y = 1 | X = x, c)}{P(Y = 1 | X = 0, c)} = RR_C(x)$

It then follows that:

$$PAF = \frac{P(Y = 1) - P(Y_0 = 1)}{P(Y = 1)}$$

$$= \frac{P(Y = 1) - E_C(P(Y = 1 | X = 0, C))}{P(Y = 1)}$$

$$= \frac{E_{X,C}(P(Y = 1 | X, C)) - E_C(P(Y = 1 | X = 0, C))}{P(Y = 1)}$$

$$= \frac{E_C(\int \pi(x | C)(P(Y = 1 | X = x, C) - P(Y = 1 | X = 0, C))dx)}{P(Y = 1)}$$

$$= \frac{E_C(\int \pi(x | C)\frac{RR(x, C) - 1}{R(x, C)}P(Y = 1 | X = x, C)dx)}{P(Y = 1)}$$

$$= \frac{\sum_c P(c)(\int \pi(x | c)\frac{RR(x, c) - 1}{R(x, c)}P(Y = 1 | X = x, c)dx)}{P(Y = 1)}$$

Now noting that:

$$\pi(x | c, Y = 1) = \frac{P(Y = 1 | X = x, c)P(c)\pi(x | c)}{P(c)P(Y = 1 | c)} = \frac{P(Y = 1 | X = x, c)\pi(x | c)}{P(Y = 1 | c)}$$

so that

$$\frac{P(c)\pi(x\,|\,c)P(Y=1\,|\,X=x,c)}{P(Y=1)} = \frac{\pi(x\,|\,c,Y=1)P(c)P(Y=1\,|\,c)}{P(Y=1)} = \pi(x\,|\,c,Y=1)P(c\,|\,Y=1)$$

, the previous displayed equation must equal:

$$= \sum_c P(c\,|\,Y=1)[\int \pi(x\,|\,C,Y=1)\frac{RR(x,c)-1}{RR(x,c)}dx]$$

$$= \int \sum_c [P(c\,|\,Y=1)\pi(x\,|\,c,Y=1)\frac{RR_C(x)-1}{RR_C(x)}]dx$$

$$= \int \pi(x\,|\,Y=1)\frac{RR_C(x)-1}{RR_C(x)}dx \qquad (1)$$

$$= E_{X|Y=1}[\frac{RR_C(X)-1}{RR_C(X)}]$$

With the second last line using Fubini's theorem to reverse the order of integration, the iterated expectation $E_{C|Y=1}\pi(x\,|\,C,Y=1) = \sum_c P(c\,|\,Y=1)\pi(x\,|\,c,Y=1) = \pi(x\,|\,Y=1)$ and

$RR(x,c) = RR_C(x)$.

**S6. Proof of generalisation of Miettinen's formula for continuous (or multi-category) exposure distributions when causal relative risks vary over confounder strata:**

The proof iqs essentially the same as the preceding proof. Note the penultimate line can be re-expressed as:

$$E_{C|Y=1}(\int \pi(x \,|\, C, Y=1)\frac{RR(x,c)-1}{RR(x,c)}dx = E_{C|Y=1}[E_{X|C,Y=1}[\frac{RR(X,C)-1}{RR(X,C)}]]$$

which is the formula in question.

### S7. Proof of formula (7)

$$\pi(x \mid Y=1)$$
$$= \frac{\pi(x)P(Y=1 \mid x)}{\int_x \pi(x)P(Y=1 \mid x)dx} \text{ (Bayes' Rule)}$$
$$= \frac{\pi(x)RR_U(x)}{\int \pi(x)RR_U(x)dx} \text{ (Divide above and below by } P(Y=1 \mid 0))$$
$$= \frac{\pi(x)RR_U(x)}{E_X(RR_U(X))}$$

Plugging the above expression into equation (1) in section S5 gives:

$$PAF = \int \frac{\pi(x)RR_U(x)}{E_X(RR_U(X))} \frac{RR_C(x)-1}{RR_C(x)} dx = \frac{E_X(RR_U(X)\frac{RR_C(X)-1}{RR_C(X)})}{E_X(RR_U(X))}$$

**S8. Analysis of the derivatives of the Relative Bias equation (4)**

$$\frac{d}{dC}\frac{1+\pi(RR_U-1)}{1+\pi(C\times RR_U-1)}\times C$$

$$=\frac{(1+\pi(RR_U-1))(1+\pi(C\times RR_U-1))-C\pi RR_U(1+\pi(RR_U-1))}{(1+\pi(C\times RR_U-1))^2}$$

$$=\frac{(1+\pi(RR_U-1))(1+\pi(C\times RR_U-1)-C\pi RR_U)}{(1+\pi(C\times RR_U-1))^2}$$

$$=\frac{(1+\pi(RR_U-1))(1-\pi)}{(1+\pi(C\times RR_U-1))^2}>\frac{(1-\pi)^2}{(1+\pi(C\times RR_U-1))^2}>0 \quad \text{for all C}$$

Derivative with respect to $\pi$:

$$\frac{d}{d\pi}\frac{1+\pi(RR_U-1)}{1+\pi(C\times RR_U-1)}\times C$$

$$=\frac{-(1+\pi(RR_U-1))(C\times RR_U-1))+(1+\pi(C\times RR_U-1))(RR_U-1)}{(1+\pi(C\times RR_u-1))^2}\times C$$

$$=\frac{RR_U(1-C)}{(1+\pi(C\times RR_u-1))^2}\times C$$

This derivative is negative for $C>1$ and positive for $C<1$

(In each case, smaller prevalences increase relative biases - but should decrease absolute biases)

As $C \to \infty$, $\dfrac{1+\pi(RR_U - 1)}{1+\pi(C \times RR_U - 1)} \times C \to 1 + \dfrac{(1-\pi)}{\pi RR_U}$

As $C \to 0$, $\dfrac{1+\pi(RR_U - 1)}{1+\pi(C \times RR_U - 1)} \times C \to 0$

**S9. Proof of Equation (10). Relative bias for a positive continuous exposure with $MREV = 0$, where $0 < 1 - \pi = P(X = 0) < 1$ and $C = \dfrac{RR_C(X)}{RR_U(X)}$ is constant for $X > 0$**

$$B = \frac{PAF_L}{PAF}$$

$$= \frac{E_X(RR_C(X)) - 1}{E_X(RR_C(X))} \frac{E_X(RR_U(X))}{E_X(RR_U(X)\frac{RR_C(X) - 1}{RR_C(X)})}$$

$$= \frac{1 - \pi + \pi E_{X|X>0}(RR_C(X)) - 1}{1 - \pi + \pi E_{X|X>0}(RR_C(X))} \frac{1 - \pi + \pi E_{X|X>0}(RR_U(X))}{\pi E_{X|X>0}(RR_U(X)\frac{RR_C(X) - 1}{RR_C(X)})}$$

$$= C \frac{1 - \pi + \pi E_{X|X>0}(RR_U(X))}{1 - \pi + \pi E_{X|X>0}(RR_C(X))}$$

$$= C \frac{1 + \pi(E_{X|X>0}(RR_U(X)) - 1)}{1 + \pi(C \times E_{X|X>0}(RR_U(X)) - 1)}$$

where the second last and last lines follow from the assumption that $RR_C(X) = C \times RR_U(X)$ when $X > 0$. The proof of equation (11) is almost identical.

**S10. Expressing,** $RR_C = \dfrac{P(Y_1 = 1)}{P(Y_0 = 1)}$**,** $RR_e = \dfrac{P(Y_1 = 1 | X = 1)}{P(Y_0 = 1 | X = 1)}$ **as weighted averages of** $RR_C(c)$**.**

Here we again assume that conditional exchangeability: $Y_x \perp X | C = c$, holds within each confounder strata $c$, for $x \in \{0,1\}$, and consistency: If $X = x$, $Y = Y_x$, for $x \in \{0,1\}$

$$RR_C = \frac{P(Y_1 = 1)}{P(Y_0 = 1)}$$

$$= \frac{E_C\{P(Y_1 = 1 | C)\}}{E_C\{P(Y_0 = 1 | C)\}}$$

$$= \frac{E_C\{P(Y_1 = 1 | X = 1,C)\}}{E_C\{P(Y_0 = 1 | X = 0,C)\}} \text{ (By exchangeability)}$$

$$= \frac{E_C\{P(Y = 1 | X = 0,C)\}}{E_C\{P(Y = 1 | X = 1,C)\}} \text{ (By consistency)}$$

$$= \frac{E_C\{(P(Y = 1 | X = 0,C)RR_C(C))\}}{E_C\{P(Y = 1 | X = 0,C)\}}$$

Similarly, we can show that

$$RR_e = \frac{E_{C|X=1}\{P(Y = 1 | X = 0,C)RR_C(C)\}}{E_{C|X=1}\{P(Y = 1 | X = 0,C)\}}$$

**Relative and absolute bias of the unadjusted Levin formula**

$$PAF = [\frac{\pi RR_U}{1+\pi(RR_U-1)}]\frac{RR_C-1}{RR_C}$$

$$PAF_{L,U} = \frac{\pi(RR_U-1)}{1+\pi(RR_U-1)}$$

$$\implies \frac{PAF_{L,U}}{PAF} = \frac{RR_C}{RR_C-1}\frac{RR_U-1}{RR_U} = C\frac{RR_U-1}{CRR_U-1}$$

**Expressing $RR_U$ in terms of $RR_C$**

$$RR_U = \frac{P(Y=1\,|\,X=1)}{P(Y=1\,|\,X=0)}$$

$$= \frac{E_{C|X=1}P(Y=1\,|\,X=1,C)}{E_{C|X=0}P(Y=1\,|\,X=0,C)}$$

$$= \frac{E_{C|X=1}(RR(C)P(Y=1\,|\,X=0,C))}{E_{C|X=0}P(Y=1\,|\,X=0,C)}$$

**Under the assumption of no effect modification, this simplifies to $RR(c) = RR_C$ for all $c$**

$$= RR_C \times \frac{E_{C|X=1}P(Y=1\,|\,X=0,C)}{E_{C|X=0}P(Y=1\,|\,X=0,C)}$$
